# Impact of ^18^FDG-avidity and immunosuppression on idiopathic and genetic cardiomyopathies

**DOI:** 10.64898/2026.01.30.26345250

**Authors:** Shiva Tabaghi, Graham H. Bevan, Stephen Hankinson, Esra D Gumuser, Mallika Lal, Madison Pico, Neal Chatterjee, Alexi Vasbinder, Richard K. Cheng, April Stempien-Otero, Neal K Lakdawala, Ron Blankstein, Marcelo F Di Carli, Benjamin Levin, Sanjay Divakaran, Babak Nazer

## Abstract

**Background:** Myocardial ^18^fluorodeoxyglucose (^18^FDG)-avidity is frequently seen in patients with genetic cardiomyopathy (CMP), as well as a growing “idiopathic ^18^FDG-avidity” group of genotype-negative patients who do not clearly have cardiac sarcoidosis (CS).

**Objectives:** To determine the prognostic implications of ^18^FDG-avidity in patients with and without genetic CMP, and the effects of immunosuppression in the latter.

**Methods:** This multicenter, retrospective study included all patients who were referred for both ^18^FDG-PET and CMP genetic testing. Patients with acute myocarditis, biopsy-proven sarcoidosis or extracardiac ^18^FDG-avidity were excluded. We investigated heart failure (HF) composite (left ventricular assist device, heart transplant, HF hospitalization, death) and arrhythmia composite (sustained ventricular arrhythmias (VT/VF), atrio-ventricular block, death) outcomes using survival analysis including Cox proportional hazards modeling and inverse probability of treatment weighting (IPWT).

**Results:** Among 372 patients, 142 (38%) were ^18^FDG-avid. Prevalence of genetic CMP among ^18^FDG-avid patients (12%) was similar to that of ^18^FDG-negative patients (19%, p=0.07). ^18^FDG-avidity was associated with increased risk of HF composite (HR 1.69 (1.04-2.75), p=0.034) and arrhythmia composite (HR 1.63 (1.1-2.4), p=0.014) outcomes compared to ^18^FDG-negative patients. However, these associations were present only in genotype-negative patients, and not in genetic CMP. After IPWT, immunosuppression of ^18^FDG-avid patients (n=49) was not associated with a reduction in HF (HR 3.31 (1.25, 8.77), p=0.016) or arrhythmia composite outcomes (HR 1.61 (0.79, 3.25), p=0.19) compared with those who were not immunosuppressed (n=93).

**Conclusions:** Myocardial-only ^18^FDG-avidity is only associated with adverse HF and arrhythmia outcomes in genotype-negative patients who do not clearly have CS. IST does not seem to modify the disease course, suggesting that not all myocardial ^18^FDG uptake reflects clinically significant inflammation.

## Introduction

Sarcoidosis is a systemic inflammatory disorder characterized by the formation of non-caseating granulomas with inflammatory infiltrate ^1,2^. Cardiac sarcoidosis (CS) is a common manifestation identified in 27%-75% of all sarcoidosis cases at autopsy^3,4^. Clinical diagnosis of *isolated* CS is particularly challenging as the yield of an endomyocardial biopsy (EMB) for detecting granulomatous inflammation is low, owing to the anatomically heterogeneous and temporally fluctuating nature of CS. The sensitivity of EMB for detecting CS is estimated between 19 and 38%^5,6^.

In 2016, the Japanese Circulation Society (JCS) released updated diagnostic guidelines for isolated CS, introducing a clinical diagnosis group that does not require histological confirmation, allowing for the diagnosis of isolated CS utilizing ^18^fluorodeoxyglucose positron emission tomography (^18^FDG-PET), cardiac magnetic resonance (CMR), and clinical presentation (ventricular arrhythmias (VT/VF), atrio-ventricular block (AVB)) with the requirement of ^18^FDG-avidity^7^. JCS criteria remains an integral component to the proposed algorithm for the diagnosis of CS in a recent Scientific Statement from the American Heart Association ^2^.

Although genetic cardiomyopathies (CMP) are a heterogenous group of diseases caused by pathogenic/likely pathogenic (P/LP) variants impacting cardiac myocyte function, the clinical presentation of genetic CMP can be similar to CS: systolic dysfunction, VT/VF, AVB. Our groups and others have previously shown that patients with P/LP variants may be suspected of having isolated CS (and often immunosuppressed) but 14-35% of patients with ^18^FDG-avidity actually have a genetic CMP ^8–19^. However, the mechanism of the ^18^FDG-avidity and the role of immunosuppression in these ^18^FDG-avid genetic CMP patients is unclear.

Additionally, as utilization of ^18^FDG-PET expands in the cardiology community, there is a growing cohort of patients with myocardial-only ^18^FDG-avidity who are genotype-negative, but who also do not clearly have isolated CS based on EMB. The mechanism of ^18^FDG-avidity and the role of immunosuppression in these “idiopathic ^18^FDG-avidity” patients is also unclear ^20^.

This multi-center retrospective cohort study was designed to evaluate the clinical features and prevalence of P/LP in patients with ^18^FDG-avid CMPs, including those meeting JCS criteria for isolated CS. Secondly, we sought to evaluate the association of ^18^FDG-avidity with adverse heart failure and arrhythmia outcomes, stratified by genetic status. In an exploratory analysis in patients with ^18^FDG-avidity, we evaluated the association of immunosuppression therapy (IST) with clinical outcomes.

## Methods

This is a multicenter retrospective cohort study including patients from the University of Washington, Oregon Health & Science University, and Brigham and Women’s Hospital which was approved by each Institutional Review Board. The study includes patients who underwent both cardiac ¹⁸FDG-PET and commercial genetic testing for genetic cardiomyopathies (Invitae Cardiomyopathy Comprehensive Panel: test code: 02251; Invitae Arrhythmia and Cardiomyopathy Comprehensive Panel: test code 02101) between 2008-2024. Variants of uncertain significance (VUS) were adjudicated by ordering providers at the sites with referral to genetics clinics if indicated, but unless they were deemed to be clinically relevant to the presentation, patients with VUS were combined with (and heretofore referred to as) genotype-negative patients with regard to further analyses. ¹⁸FDG-PET imaging was performed to assess active myocardial inflammation following standard protocols ^21^. Patients followed a high-fat, low-carbohydrate diet for at least 24 hours and fasted for 12–18 hours before imaging to suppress physiologic myocardial uptake. ¹⁸FDG-PET interpretation was done at study sites per clinical routine with ¹⁸FDG avidity (and its comparison between serial PET scans) assessed qualitatively, but not with routine use of quantitative measures such as SUV and SUV_max_.

All 3 centers followed similar clinical protocols and referred patients with suspected inflammatory or arrhythmogenic CMP for both ^18^FDG-PET imaging and genetic testing. Cardiac ¹⁸FDG-PET scans were reviewed for all patients and assessed for abnormal ¹⁸FDG uptake. Patients were classified as ¹⁸FDG-positive if at least one ^18^FDG-PET scan showed abnormal myocardial uptake. All scans with suspected incomplete myocardial ¹⁸FDG suppression (based upon clinician review) were excluded. A subset of patients was referred for EMB if ^18^FDG-avid per physician discretion, and with voltage-guidance when able^22^. Exclusion criteria included acute myocarditis (based on ACC Expert Consensus diagnostic criteria including Lake Louise MRI criteria^23,24^), biopsy-confirmed extracardiac or cardiac sarcoidosis, a clinical diagnosis of extracardiac sarcoidosis, ischemic CMP and congenital heart disease. Clinical features suggestive of extracardiac sarcoidosis meeting exclusion criteria include hilar/mediastinal lymphadenopathy, lung CT with peribronchial thickening, elevated CD4/CD8 ratio on bronchoalveolar lavage^25^.

According to the JCS criteria^7^, a diagnosis of isolated CS can be made in the absence of extracardiac involvement and biopsy confirmation when abnormal myocardial ¹⁸FDG uptake is present along with at least three additional major criteria: sustained VT/VF, high-grade AVB, abnormal wall thinning or regional wall motion abnormality on echocardiography, late gadolinium enhancement (LGE) and/or LVEF <50%. Patients meeting these criteria were considered JCS-positive.^7^

We defined two primary composite endpoints: heart failure (HF) and arrhythmic outcomes which were adjudicated across all 3 study sites. The HF endpoint was a composite of left ventricular assist device (LVAD) implantation, orthotopic heart transplant (OHT), all-cause mortality, or HF hospitalization. The arrhythmic endpoint included sustained VT or VF, new AVB requiring pacemaker implantation, and all-cause mortality with the index ¹⁸FDG-PET scan marking the beginning of the follow-up period. Outcomes were determined through review of the electronic medical record with adjudication by multiple co-authors as necessary.

Patient characteristics were summarized as counts and percentages for categorical variables and as means with standard deviations (SDs) for normally distributed continuous variables (median with inter-quartile range when not normally distributed). Chi-square and two-tailed t-tests were performed to compare the prevalence of P/LP variants versus non-P/LP results (VUS or negative) and to assess differences in baseline characteristics between patients with and without ¹⁸FDG-PET uptake. A two-sided p-value <0.05 was considered statistically significant. Time-to-event outcomes were evaluated using Cox proportional hazards models and displayed using Kaplan–Meier (KM) plots.

While our study criteria excluded patients with any systemic or biopsy-proven CS, or even any extracardiac ^18^FDG-avidity, some of our ^18^FDG-avid patients may, indeed, have had isolated CS. Thus, to further eliminate any isolated CS which may have contaminated our data, we further restricted our analyses to compare the ^18^FDG-negative group with only the ^18^FDG-avid patients who had undergone EMB (which, by inclusion, criteria were not diagnostic for any inflammatory CMP).

To evaluate the effect of IST on ^18^FDG -avidity, we compared first ^18^FDG-avid PET scan to the most recent available PET scan. To account for potential selection bias related to the deployment of immunosuppression therapy (IST), we conducted an inverse probability of treatment weighting (IPTW) analysis restricted to individuals with complete covariate data. Propensity scores were estimated using logistic regression with age, sex, race, left ventricular ejection fraction (LVEF), wall motion abnormalities, late gadolinium enhancement (LGE), premature ventricular contractions (PVC), ventricular tachycardia (VT), prior cardiac arrest, and high-grade atrioventricular block (AVB) as predictors. Stabilized IPTW weights were calculated and truncated at the 1st and 99th percentiles to reduce the influence of extreme values. Weighted KM estimators were used to obtain marginal survival curves for the primary outcomes, and weighted Cox proportional hazards models with robust variance estimators were used to derive hazard ratios and 95% confidence intervals. Covariate balance before and after weighting was evaluated using standardized mean differences. All analyses were performed in R version 4.3.1 (R Foundation for Statistical Computing, Vienna, Austria).

## Result

### Baseline Characteristics and Predictors of ^18^FDG-avidity

We identified 459 patients who had undergone both ¹⁸FDG-PET and genetic testing. Of these, 49 had biopsy-proven sarcoidosis or extracardiac ^18^FDG-avidity, and 31 incomplete myocardial ¹⁸FDG suppression and 7 patients with acute myocarditis were excluded, leaving 372 for further analysis (Fig 1). Myocardial-only ¹⁸FDG avidity was observed in 142 patients (38%). Patients with ^18^FDG-avidity were older (57±13 vs 53±14 years, p=0.005), more likely to have LGE on CMR (88 vs 71%, p=0.001), and more likely to have perfusion defects on PET (74 vs 39%, p<0.0001) than ^18^FDG-negative patients (Table 1). Patients’ initial clinical presentations and indications for testing were a mix of VT/VF, new-onset systolic HF, cardiac arrest, frequent PVCs, and high grade AVB, with many patients experiencing multiple signs or symptoms (Table 1). EMB had been performed in 44 (31%) of ^18^FDG-avid patients, most voltage-guided, and 6 from the LV. Per inclusion criteria, none of the EMBs demonstrated granulomas or inflammatory infiltrates.

**Figure 1:**
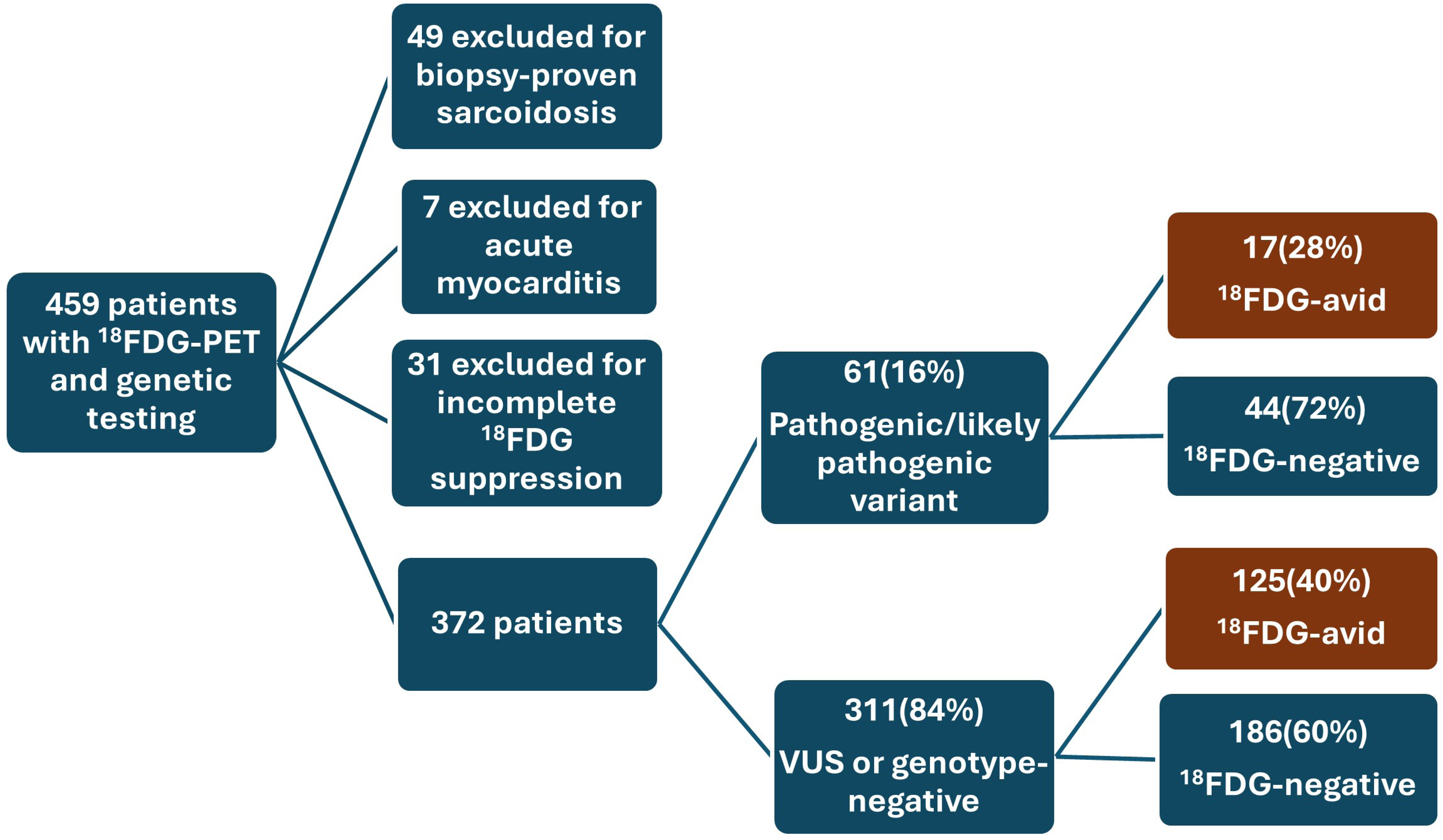
Consort Diagram of ^18^FDG-PET and genetic testing results. Abbreviations: ^18^FDG-PET: ^18^Fluorodeoxyglucose positron emission tomography VUS: Variant of Uncertain Significance

**Table 1:**

|  |  | <sup>18</sup>FDG-avid<br>(n=142) | <sup>18</sup>FDG-negative<br>(n=230) | P-value | P/LP<br>(N:61) | VUS/Negative<br>(N:311) | P-value |
| --- | --- | --- | --- | --- | --- | --- | --- |
| **Demographics** | **Age (years (SD))** | 57 (13) | 53(14) | 0.005 | 52(13) | 55(14) | 0.2 |
|  | **Gender (Female)** | 44(31%) | 84 (36%) | 0.3 | 18(29%) | 110(35%) | 0.4 |
|  | **Race (White)** | 118(83%) | 189 (82%) | 0.8 | 53(87%) | 254(82%) | 0.3 |
| **Imaging Finding** | **LGE on CMR\*** | 98(88%) | 128 (71%) | 0.001 | 34(83%) | 192(77%) | 0.4 |
|  | **LVEF (% (SD))** | 44% (14) | 46% (14) | 0.1 | 43% (13) | 46% (14) | 0.1 |
|  | **Perfusion abnormality\*\*** | 99(74%) | 79(39%) | <0.0001 | 33(62%) | 145(51%) | 0.1 |
|  | **WMA** | 53(37%) | 73(32%) | 0.2 | 20(33%) | 106(34%) | 0.8 |
| **Clinical Presentation** | **Sustained VT** | 60(42%) | 86(33%) | 0.3 | 29(47%) | 117(38%) | 0.1 |
|  | **Cardiac Arrest** | 29(20%) | 39(16%) | 0.4 | 12(20%) | 56(18%) | 0.8 |
|  | **High PVC burden** | 35(25%) | 55(23%) | 0.9 | 18(29%) | 72(23%) | 0.3 |
|  | **High grade AVB** | 28(20%) | 32(14%) | 0.1 | 11(18%) | 49(16%) | 0.6 |
|  | **New-onset HF** | 93(65%) | 136(60%) | 0.2 | 43(70%) | 186(60%) | 0.1 |
| **Endomyocardial biopsy performed** |  | 44(31%) |  |  |  |  |  |

\*: LGE was assessed in 112/142 (79%) <sup>18</sup>FDG-avid and 180/230 (78%) <sup>18</sup>FDG-negative patients, and in 251/311 (81%) VUS/negative and 41/61 (67%) P/LP patients who also underwent CMR; LGE data were missing in 22% overall.
\*\*: Perfusion abnormality was evaluated in 133/142 (91%) <sup>18</sup>FDG-avid and 202/230 (88%) <sup>18</sup>FDG-negative patients, and in 282/311 (91%) VUS/negative and 53/61 (87%) P/LP patients; perfusion data were missing in 10% overall due to limited PET perfusion tracer availability
Abbreviations:
<sup>18</sup>FDG: 18-Fluorodeoxyglucose
VUS: Variant of Uncertain Significance
P/LP: Pathogenic / Likely Pathogenic
SD: Standard Deviation
WMA: Wall Motion Abnormality
LVEF: Left Ventricular Ejection Fraction
LGE: Late Gadolinium Enhancement
CMR: Cardiac Magnetic Resonance
VT: Ventricular Tachycardia
PVC: Premature Ventricular Contraction
AVB: Atrioventricular Block

### Prevalence and Characteristics of ^18^FDG-avid Genetic Cardiomyopathies

Among ¹⁸FDG-avid patients, 12% (17/142) carried a P/LP variant, compared with 19% (44/230) of ¹⁸FDG-negative patients (p=0.07; Fig 1). Of the 142 ¹⁸FDG-avid patients, 60 (42%) met JCS criteria for isolated CS, of whom 8 (13%) were found to have a P/LP consistent with genetic CMP. JCS-positive and JCS-negative groups had similar rates of LP/P variants (13% vs 10%, p=0.7). Fig. S1

Several genes with P/LP variants were identified in ¹⁸FDG-avid genetic CMP; most commonly *LMNA*, *TTN*, and *DSP*, which represented 18%, 29% and 12% of our ¹⁸FDG-avid genetic CMP patients, respectively. No single P/LP variant was more prevalent in the ¹⁸FDG-avid compared to the ¹⁸FDG-negative groups (Table 2).

**Table 2.** Abbreviations: ^18^FDG: 18-Fluorodeoxyglucose

| Pathogenic or Likely Pathogenic Variants | <sup>18</sup>FDG-avid (n=17) | <sup>18</sup>FDG-negative (n=44) | All patients (n=61) |
| --- | --- | --- | --- |
| LMNA | 3(18%) | 11(25%) | 14(23%) |
| TTN | 5(29%) | 8(18%) | 13(21%) |
| DSP | 2(12%) | 11(25%) | 13(21%) |
| MYH7 | 2(12%) | 3(7%) | 5(8%) |
| PKP2 | 1(6%) | 3(7%) | 4(7%) |
| FLNC | 1(6%) | 2(4%) | 3(5%) |
| PLN | 1(6%) | 1(2%) | 2(3%) |
| ABCC9 | 1(6%) | 1(2%) | 2(3%) |
| DSC2 | 1(6%) | 1(2%) | 2(3%) |
| MYBPC3 | 1(6%) | 1(2%) | 2(3%) |
| DES | 2(12%) | 0 | 2(3%) |
| EYA4 | 1(6%) | 0 | 1(2%) |
| TPM1 | 0 | 1(2%) | 1(2%) |
| SCN5A | 0 | 1(2%) | 1(2%) |
| TNNI3 | 1(5%) | 0 | 1(2%) |
| RYR2 | 0 | 1(2%) | 1(2%) |
| DSG2 | 0 | 1(2%) | 1(2%) |
| TTR | 0 | 1(2%) | 1(2%) |
| CDH2 | 0 | 1(2%) | 1(2%) |
| PTPN11 | 0 | 1(2%) | 1(2%) |

### Association of ^18^FDG-avidity and genotype with clinical outcomes

Over a median[IQR] follow-up of 3.3[1.5,5.6] years, ¹⁸FDG-avidity was associated with an increased incidence for both the composite HF outcome (9 vs 5% annual incidence per patient; HR 1.69 (1.04-2.75), p=0.034; Fig. 2A) and arrhythmia composite outcomes (12 vs 7% annual incidence per patient; HR 1.63 (1.1-2.4), p=0.014; Fig. 3A) in the overall cohort. These effects were mostly driven by HF hospitalization (experienced by 15% with vs 8% of patients without ^18^FDG-avidity, p=0.02), and by sustained VT/VF (experienced by 30% with vs 17% of patients without ^18^FDG-avidity, p=0.005) for the arrhythmic composite. Incidence of death, OHT and LVAD were comparable between groups (Table 3).

**Figure 2:**
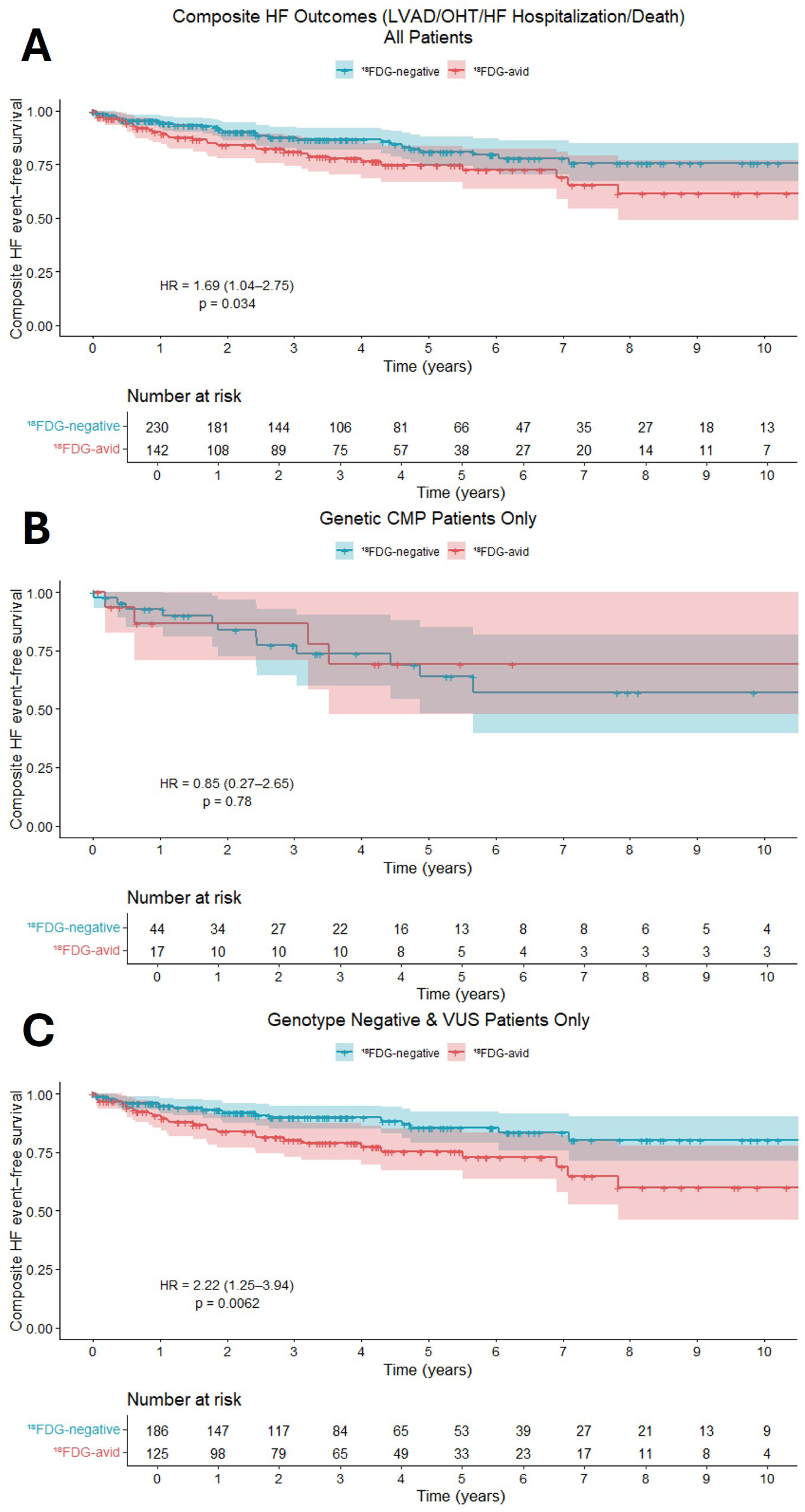
Kaplan-Meier survival analyses comparing heart failure composite outcome (LVAD, OHT, HF hospitalization and all-cause mortality) among patients with and without ^18^FDG-avidity among our entire cohort (A), genetic CMP patients (B), and genotype-negative patients (C). Abbreviations: ¹⁸FDG: 18-Fluorodeoxyglucose LVAD: Left Ventricular Assist Device OHT: Orthotopic Heart Transplant HF: Heart Failure VUS: Variant of Uncertain Significance

**Figure 3:**
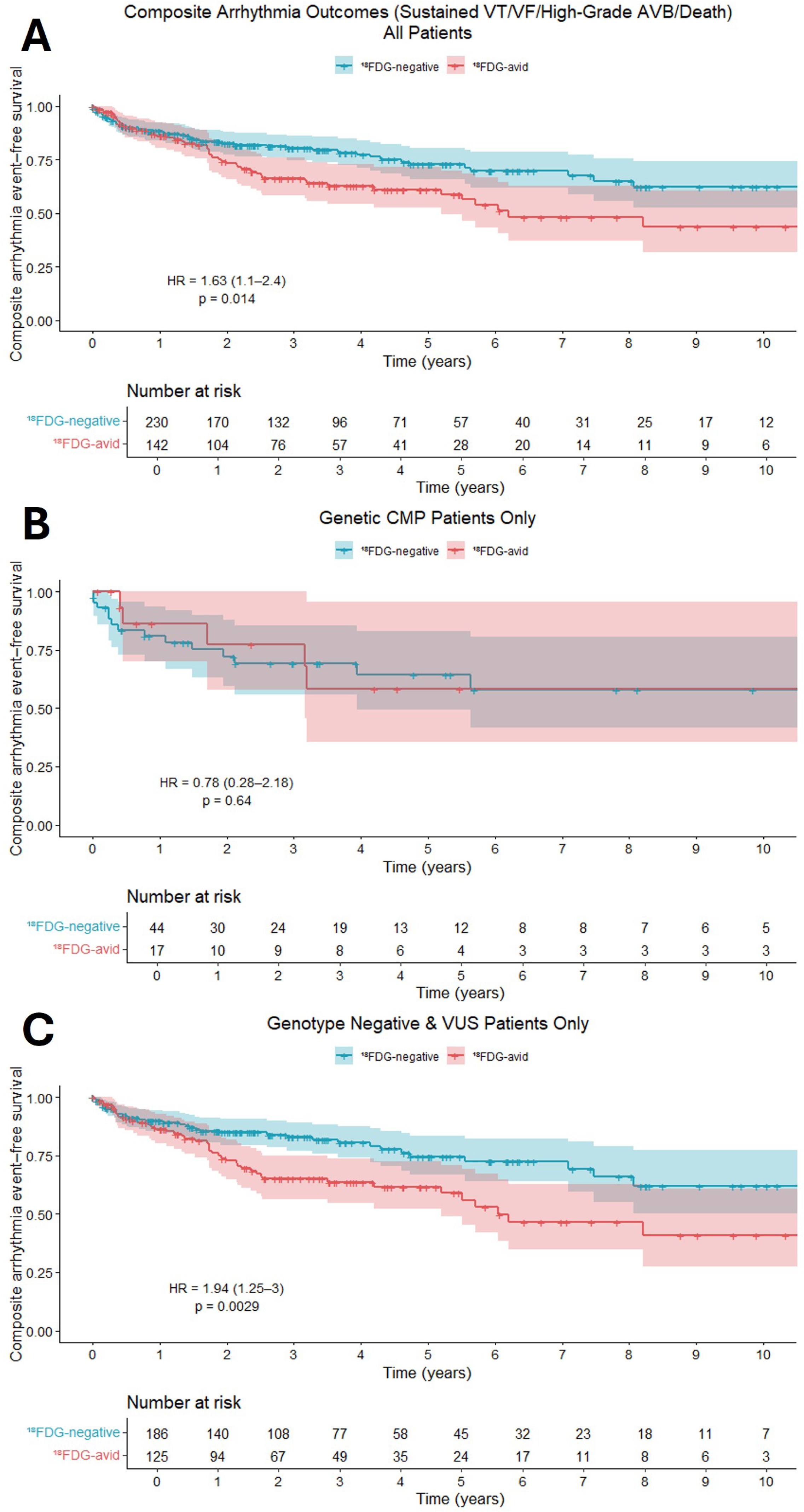
Kaplan-Meier survival analyses comparing arrhythmic composite outcome (sustained VT/VF, AV block and all-cause mortality) among patients with and without ^18^FDG-avidity among our entire cohort (A), genetic CMP patients (B), and genotype-negative patients (C). Abbreviations ¹⁸FDG: 18-Fluorodeoxyglucose VT: Ventricular Tachycardia VF: Ventricular Fibrillation AVB: Atrioventricular Block VUS: Variant of Uncertain Significance

**Table 3.**

| **Outcomes** | **<sup>18</sup>FDG-avid**<br>**(n=142)** | **<sup>18</sup>FDG-negative**<br>**(n=230)** |
| --- | --- | --- |
| LVAD/OHT | 15(11%) | 13(6%) |
| All-cause mortality | 10(7%) | 13(6%) |
| High grade AVB | 1(0.7%) | 4(2%) |
| HF hospitalization | 22(15%) | 18(8%) |
| Sustained VT/VF | 42(30%) | 40(17%) |

Abbreviations:
<sup>18</sup>FDG: 18-Fluorodeoxyglucose
LVAD: Left Ventricular Assist Device
OHT: Orthotopic Heart Transplant
AVB: Atrioventricular Block
HF: Heart Failure
VT: Ventricular Tachycardia
VF: Ventricular Fibrillation

In secondary analysis, we investigated the association of ¹⁸FDG-avidity and our two primary clinical outcomes, stratified by genetic status. Among patients with genetic CMP, ^18^FDG-avidity was not associated with a different incidence of the HF composite (HR 0.85 (0.27-2.65), p=0.78; Fig. 2B) or the arrhythmic composite (HR 0.78 (0.28-2.18), p=0.64; Fig. 3B) outcomes. By comparison, ^18^FDG-avidity was associated with an increased incidence of both outcomes among genotype-negative patients (Fig. 2C and 3C). Among this group, ^18^FDG- avid patients were more likely to meet the HF composite (HR 2.22 (1.25-3.94), p=0.006) and the arrhythmic composite (HR 1.94 (1.25-3.0), p=0.003).

In sensitivity analysis restricted to ^18^FDG-negative patients and only ^18^FDG-positive patients with negative EMB, ^18^FDG avidity was similarly associated with an increased incidence of adverse composite HF and arrhythmia outcomes (Fig. S2).

### Effects of immunosuppression on ^18^FDG-avidity and clinical outcomes

Of 142 ^18^FDG-avid patients, 49 received IST after their index PET scan and 93 did not IST agents included corticosteroids 47 (96%), Methotrexate 22 (45%) mycophenolate mofetil 10 (20%), azathioprine 3 (6%), and TNF-α inhibitor including infliximab and adalimumab 9 (18%). Patients receiving IST were younger, had slightly higher ejection fractions, and were more likely to have initially presented with sustained VT (59 vs 33%, p=0.003), but other baseline characteristics were balanced (Table 4).

**Table 4.**

|  |  | **With IST**<br>**(n=49)** | **Without IST**<br>**(n=93)** | **P-value** |
| --- | --- | --- | --- | --- |
| **Demographics** | **Age (years)** | 55(13) | 57(13) | 0.5 |
|  | **Gender (Female)** | 15(31%) | 29(31%) | 0.9 |
|  | **Race (White)** | 39(80%) | 79(85%) | 0.4 |
| **Imaging**<br>**Finding** | **LGE on CMR\*** | 37(90%) | 61(84%) | 0.3 |
|  | **LVEF (% (SD))** | 45% (15) | 43% (14) | 0.5 |
|  | **Perfusion abnormality\*\*** | 34(79%) | 65(72%) | 0.5 |
|  | **WMA** | 23(47%) | 30(32%) | 0.08 |
| **Clinical**<br>**Presentation** | **Sustained VT** | 29(59%) | 31(33%) | 0.003 |
|  | **Cardiac Arrest** | 9(18%) | 20(21%) | 0.7 |
|  | **PVC burden** | 11(22%) | 24(26%) | 0.4 |
|  | **High grade AVB** | 11(22%) | 17(18%) | 0.5 |
|  | **New-onset HF** | 32(65%) | 62(67%) | 0.9 |
| **Endomyocardial Biopsy preformed** |  | 19(39%) | 25(27%) | 0.1 |

\*: LGE was assessed in 41/49 with IST (84%) and 73/95 without IST (77%) patients who also underwent CMR: LGE data were missing in 20% overall.
\*\* : Perfusion abnormality was evaluated in 43/49 (88%) With IST and 90/93 (97%) without IST patients; perfusion data were missing in 7% overall due to limited PET perfusion tracer availability
Abbreviations:
IST: Immunosuppression Therapy
CMR: Cardiac Magnetic Resonance
LGE: Late Gadolinium Enhancement
SD: Standard Deviation
WMA: Wall Motion Abnormality
LVE: Left Ventricular Ejection Fraction
VT: Ventricular Tachycardia
PVC: Premature Ventricular Contraction

Of our 142 ^18^FDG-avid patients, 64 had at least one subsequent PET scan for comparison. When comparing these patients’ latest PET scans to their index one, both IST and no-IST groups demonstrated similarly frequent qualitative decreases in ^18^FDG-avidity (71% for both with IST and without), and similarly frequent increases in ^18^FDG-avidity (19% with IST and 8% without), with no clear impact of IST on ^18^FDG-avidity (p=0.3, Table. S1).

In an exploratory analysis, we evaluated the association of IST with our primary clinical outcomes (HF, arrhythmia) in ^18^FDG-avid patients. After IPTW-adjusted analysis controlling for the baseline differences above, ^18^FDG-avid patients treated with IST had similar incidences of HF (HR 3.31 (1.25, 8.77), p=0.016) and arrhythmia composite outcomes (HR 1.61 (0.79, 3.25), p=0.19) compared with those who were not (Fig. 4).

**Figure 4:**
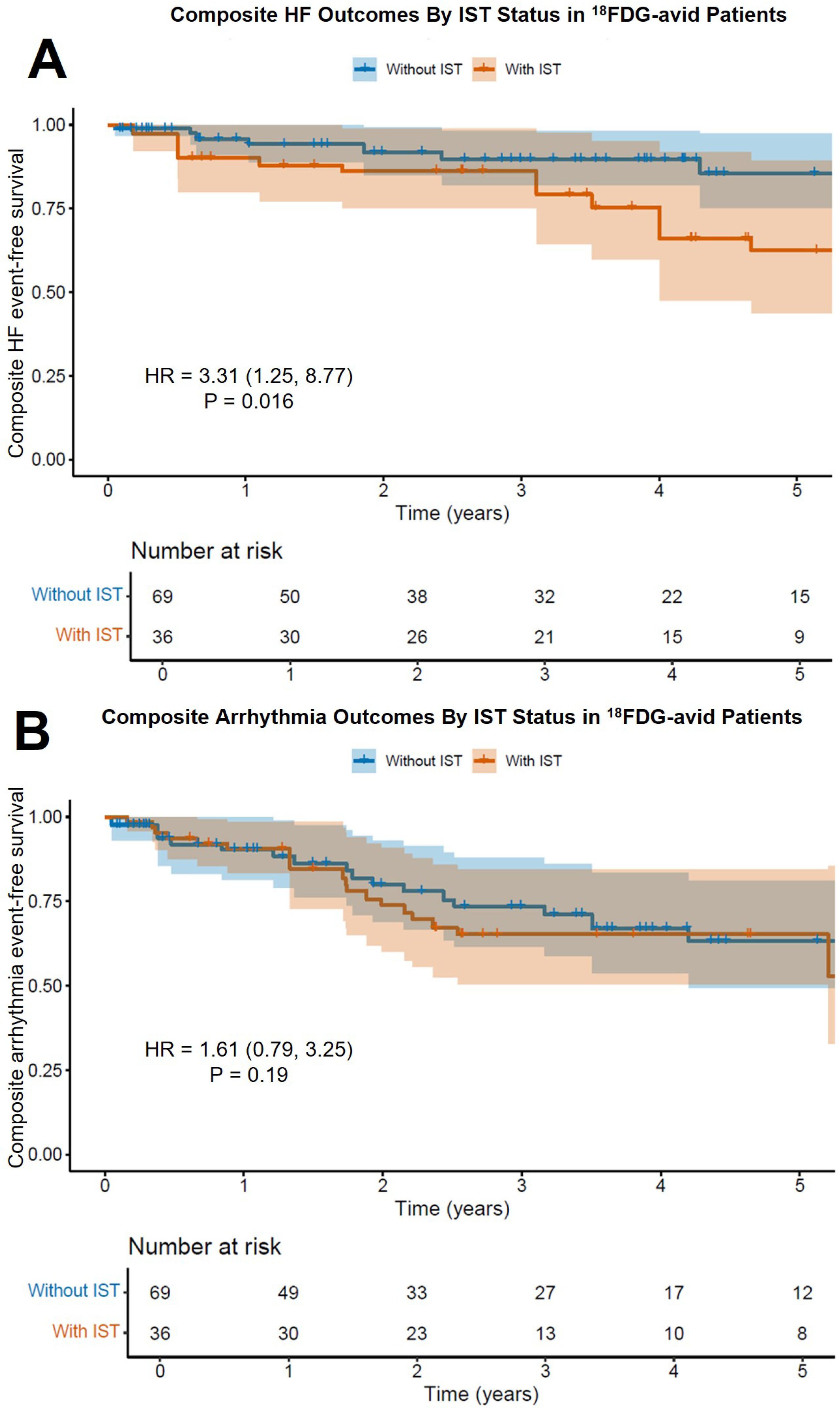
Inverse probability of treatment weighting-controlled survival analyses of ^18^FDG-avid patients receiving and not receiving IST for HF (A) and arrhythmic (B) composite outcomes. Abbreviations: IST: Immunosuppression Therapy ¹⁸FDG: 18-Fluorodeoxyglucose HF: Heart Failure LVAD: Left Ventricular Assist Device OHT: Orthotopic Heart Transplant AVB: Atrioventricular Block VT: Ventricular Tachycardia VF: Ventricular Fibrillation AVB: Atrioventricular Block HF: Heart Failure

Given the clinical and research methodological challenges in definitively excluding isolated CS, we studied the effects of IST exclusively among the ^18^FDG-avid patients who would have met the JCS criteria for isolated CS. Even in this group, IST was not associated with a reduction in HF or arrhythmic outcomes (Fig. S3).

## Discussion

Prior work by our groups and others has identified ¹⁸FDG-avidity in genetic CMP including P/LP variants associated with arrhythmia (e.g. DSP, PKP2), hypertrophic cardiomyopathy (e.g. HCM, MYH7, MYBC3), and other genetic causes (e.g. LMNA, TTN, FLNC)^8,9,11,12^. The present study builds upon this work with a large cohort of genotyped and ^18^FDG-phenotyped patients, reinforcing that genetic CMP can masquerade as isolated CS, with 12% of ^18^FDG-avid patients and 13% of patients who met JCS diagnostic criteria for isolated CS harboring a P/LP variant consistent with genetic CMP. While we identified a broad array of genes (Table 2) implicated in this phenomenon, none demonstrated a specific predilection for or against ^18^FDG-avidity. Our findings suggest that contemporary diagnostic guidelines for isolated CS that do not require a tissue diagnosis may lack specificity, and further that the diagnosis of isolated CS should occur only after endomyocardial biopsy and excluding genetic CMP.

### Insight into mechanisms of ^18^FDG-avidity

Our study suggests that among patients with genetic CMP, ^18^FDG-avidity was not associated with an increased risk of adverse HF or arrhythmic outcomes calling into question whether the ^18^FDG-avidity represents true (or at least clinically relevant) inflammation. When phenotyping patients with non-ischemic CMP, ^18^FDG-avidity is typically thought to represent inflammation. However, non-specific ^18^FDG -avidity is common^20,26^, and it should be noted that a long-standing use of ^18^FDG-PET scans in the field of coronary artery disease (CAD) is the “viability study” for the detection of “viable” myocardium which shifts its metabolism from fatty acids to glucose, and becomes ^18^FDG-avid in the setting of long-standing ischemia^27^. We hypothesize that genetic CMPs similarly shift myocardial metabolism due to impaired sarcomeric, cytoskeletal or mitochondrial function. Aoyama, et al. systematically performed ^18^FDG-PET scans before and after septal reduction therapies (SRT) in HCM patients, and demonstrated that ^18^FDG-avidity was markedly (and diffusely) reduced post-SRT, a finding that lends itself to more of a metabolic than inflammatory mechanism^9^. Our study’s findings further support this as we demonstrate that many patients’ (85%) ^18^FDG-avidity decreased spontaneously (without IST) on subsequent PET scans, and that IST was not associated with a reduction in ^18^FDG-avidity.

There is some evidence to the contrary of our hypothesis. Mouse models of genetic CMP have been shown to have some myocardial inflammation, but this appears to be mostly driven by a macrophage infiltration in response to genetically-mediated myocardial injury^28^. Peretto et al.^12^ performed EMB of genetic CMP patients with ^18^FDG avidity and found lymphocytic infiltrates consistent with myocarditis in 16% (4 of 25 patients). The Peretto study used sensitive criteria for inflammatory cell infiltrates which may have differed from the clinical methods and thresholds used by pathologists evaluating biopsy specimens in this study. It should also be noted that many of the patients in the Peretto study were biopsied during “hot phase” (acute myocarditis presentations which have been described in patients with genetic CMP, and which we systematically excluded). Indeed, patients presenting with acute myocarditis have been shown to have a higher prevalence of pathogenic variants (8%) than healthy controls (<1%)^11^, but this may represent a “two-hit hypothesis” in which an underlying desmosomal, sarcomeric or other myocyte genetic vulnerability renders genetic CMP patients more vulnerable to a more aggressive course of viral CMP^14^. We excluded acute myocarditis, our PET scans were performed as part of diagnostic evaluation for arrhythmias and/or CMP (Table 1), and none of our 44 EMBs demonstrated inflammatory infiltrates. Additionally, among other recent studies and case series of ^18^FDG-avid genetic CMPs, none of the 36 EMBs in these studies demonstrated inflammatory infiltrates^8,15–19^. It is well-described that the failing and remodeling heart shifts its fuel source (as in PET viability studies for CAD) from free fatty acids to glucose^29–31^.

Ultimately, as we excluded “hot phase” myocarditis presentations and biopsy-proven CS, our findings suggest that not all ^18^FDG-avidity seen clinically arises from a primary inflammatory process, but either from a secondary immune response to myocyte injury or a metabolic switch from free fatty acids to preferential gluconeogenesis in myocardium.

### Diagnostic and Therapeutic Quandaries of Myocardial-Only ^18^FDG-avidity

One meaningful limitation of our study is that among our genotype-negative group, even though we excluded patients with systemic sarcoid or any extracardiac ^18^FDG-avidity, we cannot definitively exclude isolated CS given the known limitations of diagnostic sensitivity of EMB and the fact that some patients did not undergo EMB. Even in the subset (31%) of our cohort that underwent EMB (none of which demonstrated inflammation or granulomas despite use of voltage-guidance and LV sampling), the anatomically heterogeneous and temporally fluctuating nature of CS could have resulted in “false negative” EMB results. However, this limitation reflects a real-world conundrum in clinical practice regarding how to classify and treat patients who are genotype-negative, EMB-negative, and ^18^FDG-avid: an “idiopathic ^18^FDG-avid CMP”.

With respect to prognostication and management, we would note that among patients with *biopsy-proven CS*, studies consistently demonstrate an association of cardiac ¹⁸FDG-avidity with adverse cardiovascular outcomes including HF, VT/VF, and death ^2,32–34^. IST in ^18^FDG-avid patients with CS is appears to be associated with improved clinical outcomes in non-randomized studies^12,35–37^. In our study, we show that among genotype-negative patients, ^18^FDG-avidity was also associated with increased incidence of HF and arrhythmic outcomes, but IST was not associated with a reduction in these outcomes. Though exploratory, our data suggests that empiric use of IST in ^18^FDG-avid patients may not improve clinical outcomes. Future efforts to evaluate the clinical benefit of IST in such patients are warranted.

### Limitations

While the study was performed at 3 high-volume nuclear medicine centers, variability in PET acquisition and interpretation in the absence of a core facility may affect our results. Similarly, our EMB data are based on clinical pathology interpretations performed at the 3 sites, and were not interpreted at a core, or using advanced methods such as immunohistochemistry or spatial transcriptomics. Since only 31% of our ¹⁸FDG-avid patients underwent EMB, we cannot exclude the possibility that some isolated CS patients were included in the ¹⁸FDG-avid cohort. However, patients with any evidence of acute myocarditis (including the “hot phase” myocarditis described in genetic CMP patients), extracardiac sarcoidosis or extracardiac ¹⁸FDG uptake were systematically excluded. In addition, we performed a sensitivity analysis comparing only ¹⁸FDG-avid patients with negative EMB to all ^18^FDG-negative patients and found similar prognostic associations of ^18^FDG avidity with clinical outcomes.

## Conclusions

Our study informs clinical care for patients found to have myocardial-only ^18^FDG-avidity in that they should:

1. undergo genetic testing, and EMB when clinically appropriate (the latter with voltage guidance and LV sampling, when able)
2. be closely monitored and treated for arrhythmias and worsening HF given the association of ^18^FDG-avidity with an increase in these types of adverse outcomes
3. assuming EMB is not diagnostic for CS, *not* be empirically immunosuppressed (while we await further data) as IST was not associated with a reduction in either ^18^FDG-avidity or HF/arrhythmic endpoints

Understanding the mechanism of ¹⁸FDG-avidity in this population may unlock novel therapeutic approaches to treatment of non-ischemic CMPs.

## Data Availability

Anonymized patient data can be requested from the corresponding author.

## Acknowledgements

The authors thank the nuclear imaging and medical genetics teams at their 3 institutions (OHSU, BWH, UW) for their high-quality care.

## Twitter

18FDG-avidity is associated with adverse HF and arrhythmia outcomes only in genotype-negative patients without clear cardiac sarcoid. In absence of biopsy-proven sarcoid, immunosuppression doesn’t modify disease courses @B_Naz_MD

## Abbreviations

CS: Cardiac sarcoidosis
EMB: Endomyocardial biopsy
JCS: Japanese Circulation Society
^18^FDG-PET: ^18^Fluorodeoxyglucose positron emission tomography
CMP: Cardiomyopathies
P/LP: Pathogenic/likely pathogenic
VUS: Variants of uncertain significance
HF: Heart failure
IST: Immunosuppression therapy
VT/VF: Ventricular arrhythmias
AVB: Atrio-ventricular block
LGE: Late gadolinium enhancement
LVAD: Left ventricular assist device
OHT: Orthotopic heart transplant

---

**Table S1.**

| Change between first and most recent <sup>18</sup>FDG-PET | With IST (n=40) | Without IST (n=24) | P-value |
| --- | --- | --- | --- |
| Increased <sup>18</sup>FDG-avidity | 7(19%) | 2(8%) | 0.3 |
| Stable <sup>18</sup>FDG-avidity | 4(10%) | 5(21%) |  |
| Decreased <sup>18</sup>FDG-avidity | 29(71%) | 17(71%) |  |

Abbreviations:
IST: Immunosuppression Therapy
<sup>18</sup>FDG: 18-Fluorodeoxyglucose

**Figure S1**

Genetic testing results among ^18^FDG-avid patients meeting JCS criteria for isolated CS.

Abbreviations:

JCS: Japanese Circulation Society

¹⁸FDG: 18-Fluorodeoxyglucose

VUS: Variant of Uncertain Significance

**Figure S2**

Kaplan-Meier survival analyses comparing HF (A) and arrhythmic (B) composite outcomes between ^18^FDG-negative patients and only the ^18^FDG-avid patients who had undergone EMB, further excluding data contamination by any isolated CS.

Abbreviations:

HF: Heart Failure

¹⁸FDG: 18-Fluorodeoxyglucose

EMB: Endomyocardial Biopsy

LVAD: Left Ventricular Assist Device

OHT: Orthotopic Heart Transplant

VT: Ventricular Tachycardia

VF: Ventricular Fibrillation

AVB: Atrioventricular Block

**Figure S3**

Inverse probability of treatment weighting-controlled survival analyses of ^18^FDG-avid patients meeting JCS criteria for isolated CS who were receiving and not receiving IST for HF (A) and arrhythmic (B) composite outcomes.

Abbreviations:

¹⁸FDG: 18-Fluorodeoxyglucose

JCS: Japanese Circulation Society

CS: Cardiac Sarcoidosis

IST: Immunosuppression Therapy

HF: Heart Failure

LVAD: Left Ventricular Assist Device

OHT: Orthotopic Heart Transplant

VT: Ventricular Tachycardia

VF: Ventricular Fibrillation

AVB: Atrioventricular Block

